# Heritability enrichment implicates microglia in Parkinson’s disease pathogenesis

**DOI:** 10.1101/2020.11.25.20238162

**Authors:** Maren Stolp Andersen, Sara Bandres-Ciga, Regina H. Reynolds, John Hardy, Mina Ryten, Lynne Krohn, Ziv Gan-Or, Inge R. Holtman, International Parkinson’s Disease Genomics Consortium, Lasse Pihlstrøm

## Abstract

**Objective:** Understanding how different parts of the immune system contribute to pathogenesis in Parkinson’s disease is a burning challenge with important therapeutic implications. We studied enrichment of common variant heritability for Parkinson’s disease stratified by immune and brain cell types.

**Methods:** We used summary statistics from the most recent meta-analysis of genome-wide association studies in Parkinson’s disease and partitioned heritability using linkage disequilibrium score regression, stratified for specific cell types as defined by open chromatin regions. We also validated enrichment results using a polygenic risk score approach and intersected disease-associated variants with epigenetic data and expression quantitative loci to nominate and explore a putative microglial locus.

**Results:** We found significant enrichment of Parkinson’s disease risk heritability in open chromatin regions of microglia and monocytes. Genomic annotations overlapped substantially between these two cell types, and only the enrichment signal for microglia remained significant in a joint model. We present evidence suggesting *P2RY12*, a key microglial gene and target for the anti-thrombotic agent clopidogrel, as the likely driver of a significant Parkinson’s disease association signal on chromosome 3.

**Interpretation:** Our results provide further support for the importance of immune mechanisms in PD pathogenesis, highlight microglial dysregulation as a contributing etiological factor and nominate a targetable microglial gene candidate as a pathogenic player. Immune processes can be modulated by therapy, with potentially important clinical implications for future treatment in Parkinson’s disease.

## Introduction

Understanding how the immune system contributes to pathogenesis is a burning challenge in Parkinson’s disease (PD) and other neurodegenerative disorders, which could have major importance for future therapy. To date, the treatment for PD is merely symptomatic, yet the prospect of immune modulation is one of the most promising disease-modifying strategies. However, immune processes are likely to have both protective and harmful influences in the context of neurodegeneration. Thus, a deeper understanding of the role of different cell types and molecular processes is urgently needed.

Microglia are the primary immune cells of the central nervous system, similar to peripheral macrophages, yet specialized for the immunoprivileged environment inside the blood-brain barrier. Neuropathological and experimental studies have demonstrated microglial activation in PD brains^1^, yet it is not clear whether this activation is part of the pathogenic process or a protective response. Furthermore, several lines of evidence have suggested a role for the peripheral immune system^2-7^, but the specific significance of different immune mechanisms for pathogenesis, their role in the causal hierarchy of the disease and the interrelation between microglial and peripheral immune processes remain poorly understood.

Genetic approaches have the great advantage that the direction of causality is generally straightforward to interpret. A signal from the HLA region was among the earliest identified susceptibility loci in genome-wide association studies (GWAS) of PD, highlighting a potential causal role of the immune system.^8^ The latest meta-analysis of PD GWAS identified 90 genome-wide significant risk variants, for which the relevant genes and mechanisms remain largely unknown.^9^ Recently, a number of powerful statistical methods have been developed to infer involvement of biological pathways and cell types in disease pathogenesis based on association data from large GWAS combined with functional annotation of the human genome.^10^

Previously published analyses of heritability enrichment have unequivocally highlighted immune mechanisms in Alzheimer’s disease (AD)^11^, implicating monocytes, macrophages and microglia as likely relevant cell types.^12^ However, similar analyses in PD have thus far yielded mixed results. While some studies have reported evidence of heritability being enriched in immune-related regions,^3, 11^ other large-scale analyses have seen significant results for brain or neurons only^13^, potentially due to methodological differences. Importantly, no study to date has reported enrichment of heritability linking genetics to microglia in PD.

We used the largest set of summary statistics from PD GWAS to date to investigate heritability enrichment partitioned by cell type, focusing on immune and brain cells. We show significant enrichment of PD heritability in open chromatin regions of microglia, an enrichment that was stronger than that observed for neurons. Significant enrichment was also observed for monocytes, but not when open chromatin regions overlapping with microglia were excluded. Combining available GWAS statistics, genomic annotations and published gene expression data to interpret an individual GWAS locus, we highlight *P2RY12* as an example of a likely microglial causal gene underlying a PD association signal.

## Methods

### Genetic association data

For stratified linkage disequilibrium score regression (s-LDSC) analyses, we used the full summary statistics from the latest meta-analysis of PD GWAS, which included a total of 37,688 cases, 18,618 proxy-cases and 1,417,791 controls.^9^ To validate the results from s-LDSC, we also performed polygenic risk score analyses, following the same approach as we applied in a recently published study.^14^ Similar to this study, we took advantage of genetic data available through the International Parkinson’s Disease Genomics Consortium (IPDGC): Summary statistics from an earlier meta-GWAS of 26,035 PD cases and 403,190 controls of European ancestry^15^ were used as a reference dataset to define risk allele weights, and individual genotypes not included in this earlier meta-GWAS were assigned to a training dataset (7,218 PD patients and 9,424 controls) and a test dataset (5,429 PD cases and 5,814 controls).

### Tissue and cell type annotations

With an interest to understand the contribution of microglia and peripheral immune cells, we primarily aimed to analyze cell-specific data from brain and immune cells. However, as previous studies have used different methods to define regions of relevance for specific tissues and cell types, we initially contrasted tissue-level approaches based on gene expression versus approaches based on open chromatin regions as assessed by Assay for Transposase-Accessible Chromatin sequencing (ATAC-seq). As an example of an expression based approach, we mirrored the method followed for data from the Genotype-Tissue Expression (GTEx) project database^16^ in a previous publication on heritability enrichment of specifically expressed genes.^17^ In that study, normalized gene expression values were compared across tissues and t-statistics were calculated for specific expression in each focal tissue. Genes were ranked by their t-statistic, and the 10% of genes with the highest t-statistic were defined as the set of specifically expressed genes corresponding to the focal tissue. We downloaded the relevant specifically expressed gene sets as provided by the authors (Alkes lab, see online resources) for the immune and central nervous systems, as well as three additional systems as negative controls (musculoskeletal, cardiovascular and gastrointestinal), merging gene sets for 21 different tissues into these five tissue categories. We used the online Ensembl tool BioMart to download b37 coordinates for all genes and added 100kb of flanking sequence, still following the method from Finucane *et al*.^17^

To define open chromatin in individual brain cell types, we used ATAC-seq peak data from neurons, oligodendrocytes, astroglia and microglia generated by Nott *et al*.^18^ The union of all these datasets was merged into a common brain ATAC-seq dataset. Similarly, ATAC-seq data from monocytes, B-cells, CD4+ and CD8+ T-cells generated in a study by Corces *et al*.^19^ were downloaded as BigWig files, peaks were called using macs2^20^ and the union of all peaks combined to form a merged set of immune cell peaks. ATAC-seq peaks from three negative control tissues were downloaded from the ENCODE Reference Epigenome^21^ (accessed Sep 11th 2020) and merged into combined open chromatin sets for gastrointestinal tissue, striated muscle and cardiovascular tissue. To visualize and analyze the degree of overlap across different tissues and annotation approaches, we used the command-line tool Intervene.^22^

### Stratified LD score regression

Linkage disequilibrium (LD) is the non-random association of alleles at different loci in a given population, as when variants located close to each other on the same chromosome tend to be inherited together as a haplotype. LD score regression is a statistical framework that takes summary statistics from GWAS as an input and examines the relationship between test statistics and LD in order to estimate common variant heritability. Combined with a set of annotations that assign genomic regions to functional categories, a stratified version of this approach (s-LDSC) can be used to infer relevant pathways or cell types showing a significant enrichment of heritability relative to the rest of the genome. We applied s-LDSC to partition heritability for PD across tissue and cell-type annotations and assessed enrichment, following the method as described by Finucane *et al*.^23^ We used downloadable LD scores estimated from 1000 Genomes data and included single-nucleotide polymorphisms (SNPs) present in HapMap 3 with an allele frequency above 0.05, excluding the Major Histocompatibility Complex region (see online resources). Enrichment of heritability was assessed controlling for the effects of 53 functional annotations included in the full baseline model v.1.2. P-values were calculated based on the coefficient *z*-score, as recommended by the authors. Enrichment of heritability linked to the brain and immune system has been reported before, and we considered enrichment passing a nominal p-value threshold (p<0.05) as replication of previous work in the initial tissue-level analysis, although we also included three other tissues as negative controls. In the analyses of brain and immune cell types we adjusted the significance threshold for eight individual cell types (p < 0.00625).

### Polygenic risk score analysis

A polygenic risk score (PRS) is a score that summarizes the effect of many alleles on an individual’s phenotype, using summary statistics from a large GWAS to determine allele weights, which are then applied to individual-level data in an independent sample set for which scores are generated. As with LDSC, the analysis can be restricted to subsets of the genome to stratify for functional pathways or cell types of interest. To validate our heritability enrichment results using an independent method, we implemented a PRS approach recently used to investigate genetic risk across curated functional pathways in PD (PMID: 32601912).^14^ PRSice v.2.3e^24^ was used to estimate individual PRS in the training and test datasets, based on allele weights from reference data summary statistics restricted to SNPs within open chromatin regions of specific brain and immune cell types, as defined by ATAC-seq peaks. PRSice allows for empirical optimization of the p-value threshold for inclusion of SNPs into the PRS calculation. However, to avoid overfitting and obtain comparable results across cell types, we set a predefined threshold of p < 0.05. We used 1000 permutations to calculate Nagelkerke’s pseudo r^2^, coefficient and empirical p-values for the association between PRS and disease status in the training and test sets, adjusting for an estimated PD prevalence of 0.005, age at onset for cases and age at examination for controls, sex, and 20 principal components to account for population stratification.

### Overlap with GWAS loci

After assessing enrichment of heritability based on genome-wide summary statistics, we wanted to explore if potentially causal SNPs at significant GWAS-loci locate specifically to microglial ATAC-seq peaks, and if a functional role in microglia could be supported by other publicly available data for any PD GWAS locus. We used the statistical fine-mapping method PAINTOR (Probabilistic Annotation INTegratOR; v3.1)^25^ to prioritize 95% credible sets at PD GWAS loci without functional annotations as priors. We excluded the *GBA* locus, where causal variants are known to be protein coding^26^, and two loci of particularly complicated LD structure, namely the *HLA* region and the H1/H2 inversion haplotype region on chromosome 17q21.31. We also merged loci that were less than 1Mb apart. For each locus in a total of 71 loci, we extracted summary statistics for all variants within a window extending 10kb from the first and last genome-wide significant SNP within the locus. PAINTOR was run according to the recommendations in the online documentation using reference data from 1000 Genomes Phase 3 without priors (see online resources). We then merged 95% credible sets from all 71 loci and assessed the overlap with ATAC-seq peaks for different cell types using bedtools. Co-localization of credible set SNPs and open chromatin was further evaluated taking advantage of publicly available databases and relevant published results.^27-31^ Overlap between credible set SNPs and genomic annotations at the *P2RY12* locus were visualized using LocusZoom and UCSC Genome Browser.

## Results

### Stratified LD score regression shows enrichment for microglia, monocytes and neurons

Previous studies of heritability enrichment in PD have varied both with respect to methods and reported results. Reviewing the literature, we noted that some studies link genomic regions to cell types based on the location of expressed genes^14, 32^, whereas others take an approach based on available epigenetic data.^18, 33^ Regions annotated based on GTEx-expressed genes span several hundred kilobases compared to narrow ATAC-seq peaks of typically a few hundred base pairs, indicating that the power to detect enrichment may be very different depending on annotation approach, in line with previous reports.^34^

Analyzing heritability enrichment on the tissue group level using s-LDSC with both annotation approaches, we found enrichment for brain tissue with both GTEx (coefficient p = 0.0038) and ATAC-seq annotations (coefficient p = 0.0072), whereas immune tissue enrichment was nominally significant only using ATAC-seq annotations (coefficient p = 0.032) (Figure 1). Cardiovascular, gastrointestinal or muscle tissue did not show enrichment with either method. We took these results as an indication that the annotation approach based on open chromatin, as a means to identify genomic regulatory elements, would be the most relevant to take forward to cell type specific enrichment analysis of brain and immune cells. We note however, that a methodological comparison of annotation approaches was not the main aim of our work, and that the methods were not identical in all other respects, e.g. starting from tissue versus cell-type level data.

**Figure 1.**
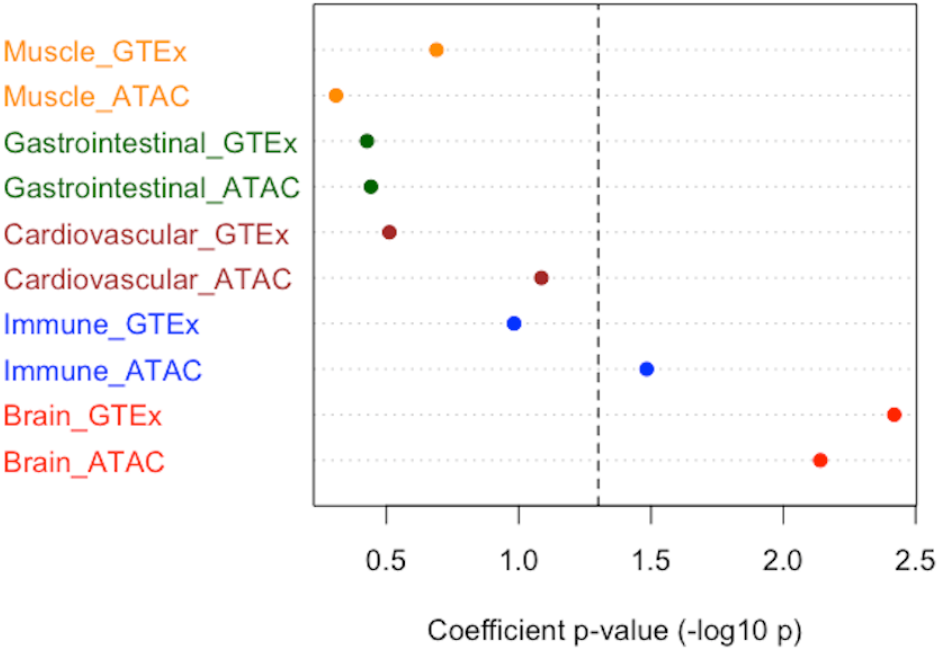
Stratified LD score regression at tissue level. Results from s-LDSC applied to five major tissue groups using either specifically expressed genes or open chormatin regions as identified by ATAC-seq to partition heritability. The dashed line represents a nominal significance threshold of one-sided p<0.05.

Assessing partitioned heritability in open chromatin regions of specific brain cell types, the strongest enrichment was seen for microglia (coefficient p = 0.0046), with nominally significant enrichment also for oligodendrocytes (coefficient p = 0.0097) and neurons (coefficient p = 0.048) (Figure 2). In immune cells, enrichment was significant for monocytes, but did not pass a corrected p-value threshold for B-cells, CD4+ or CD8+ T-cells. Calculating the pairwise Jaccard statistics between brain and immune cell pairs confirmed that microglia shared more open chromatin with immune cells than with other brain cell types, and most of all with monocytes (Figure 3 and 4). To further explore whether enrichment signals for microglia and monocytes are independent, we followed a previously reported strategy^34^ and jointly analyzed microglia and monocyte annotations in the same model, conditional on the baseline annotations. In a joint model, the microglia enrichment signal retained nominal significance (coefficient p = 0.037), whereas monocytes did not (coefficient p = 0.26).

**Figure 2.**
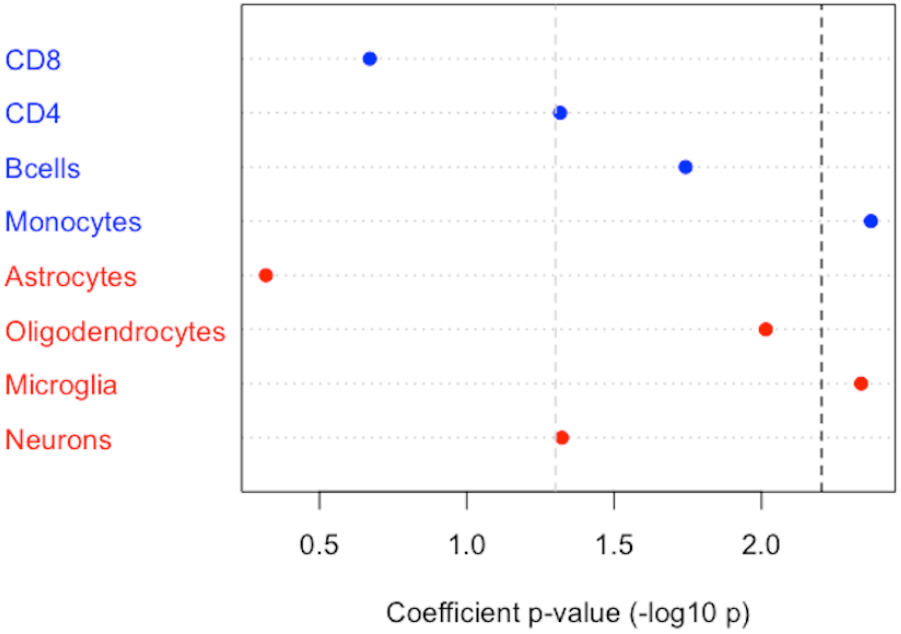
Stratified LD score regression in immune and brain cells. Results from s-LDSC applied to immune (blue) and brain cells (red) using open chormatin regions as identified by ATAC-seq to partition the genome. The dashed line represents a Bonferroni-corrected significance threshold of one-sided p<0.00625.

**Figure 3.**
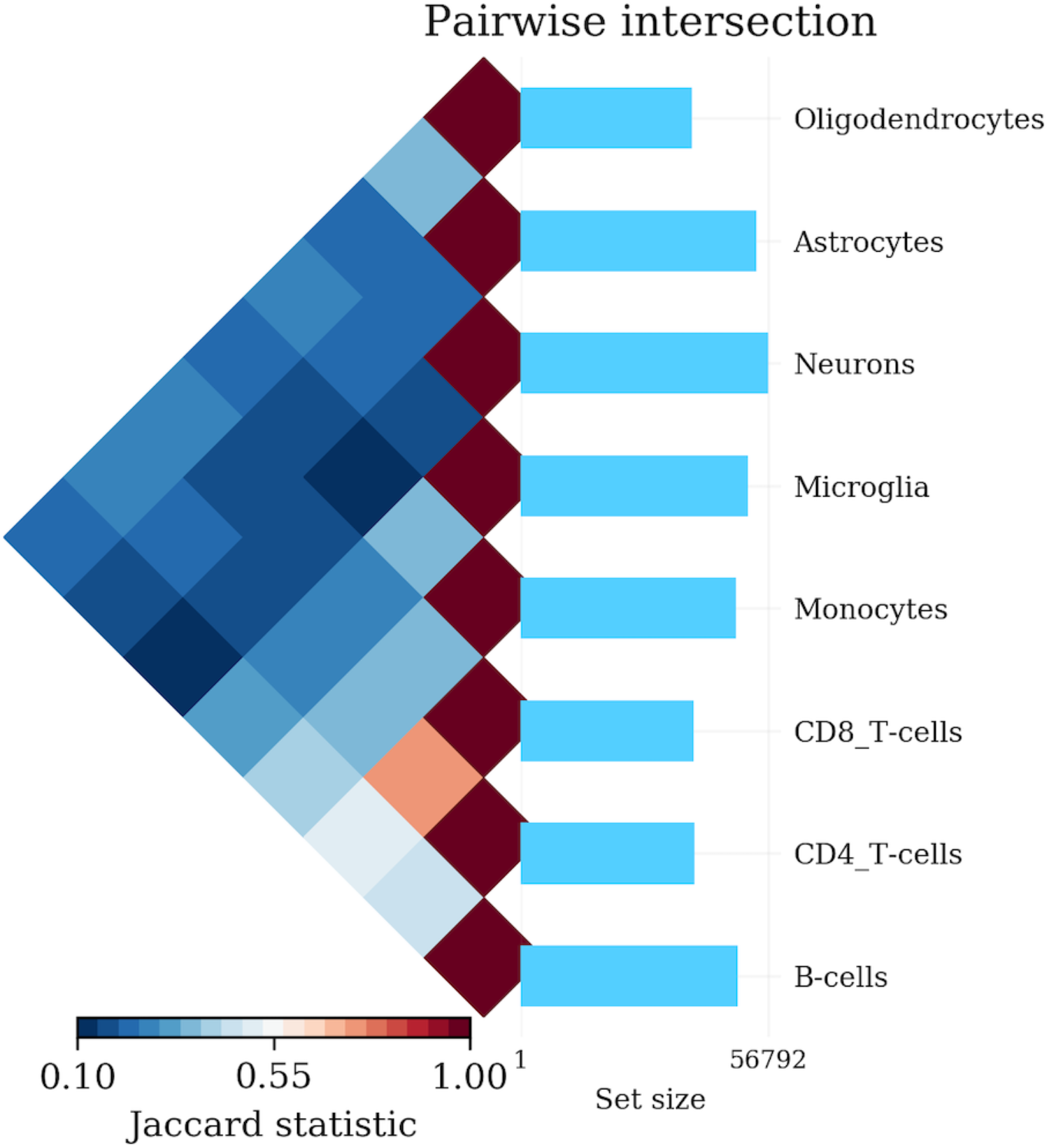
Pairwise Jaccard statistics for brain and immune cell ATAC-seq peaks. The plot shows Jaccard statistics for the pairwise intersection of ATAC-seq peaks for investigated immune and brain cell types (left) and the total size of open chormatin regions for each cell type (right). Microglia show more overlap with immune cells than other brain cells, and highest overlap with monocytes.

**Figure 4.**
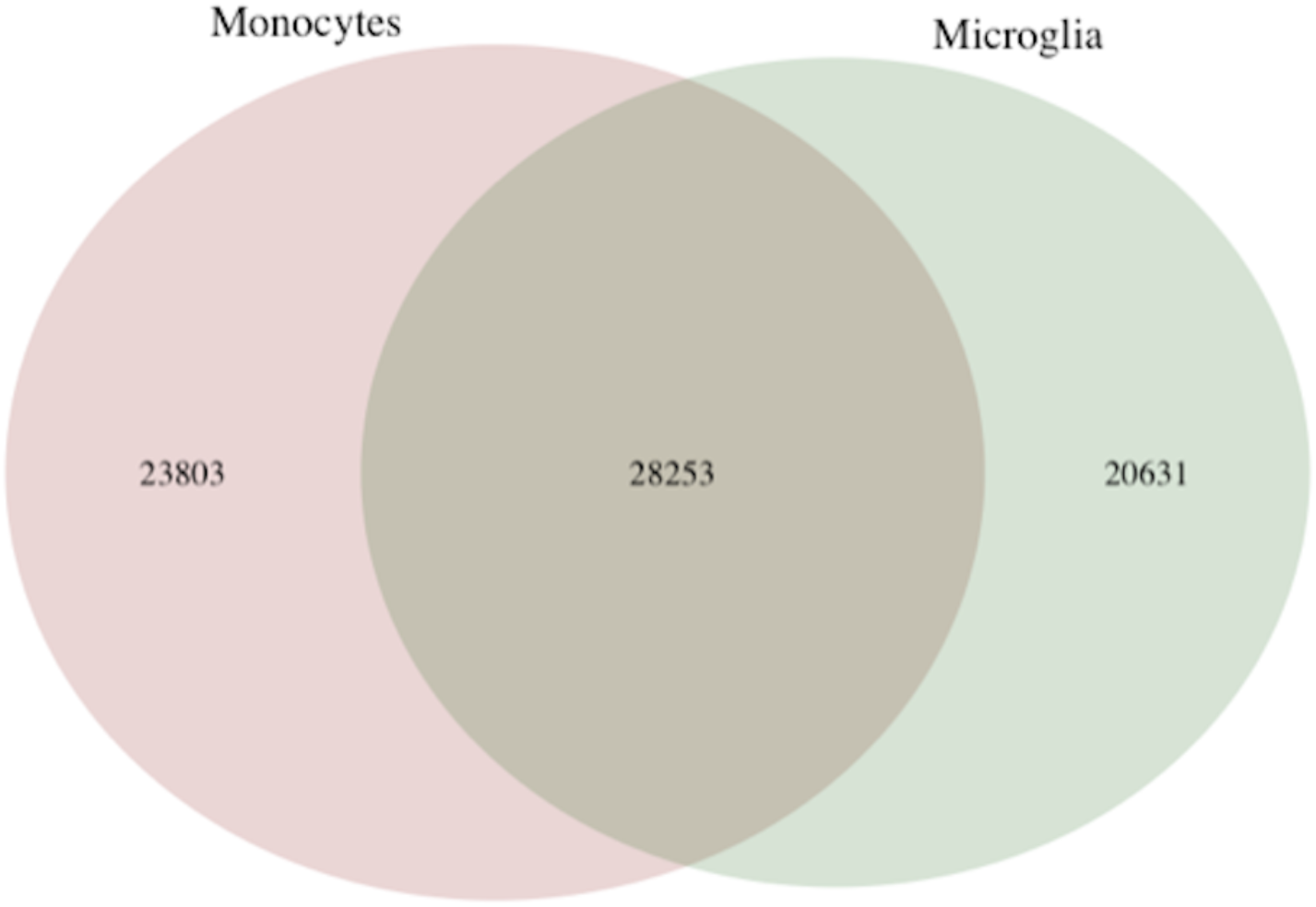
Venn diagram of genomic positions within ATAC-seq peaks of microglia and monocytes. Venn diagram showing the proportion of overlapping versus unique open chromatin as assessed by ATAC-seq in monocytes and microglia.

### A polygenic risk score approach provides further support for microglial enrichment

Investigating the performance of PRS calculated across open chromatin regions we expectedly found very strong p-values for all cell types. We note that this method is far less conservative, as it does not control for other co-localizing annotations in the same way as the baseline model in s-LDSC. Of interest, however, is the relative performance of each cell-specific PRS. In line with the results from s-LDSC, we found that PRS estimates based on SNPs in microglial open chromatin regions showed the strongest association with PD of all the brain cell types in both the training and test datasets (Table 1). In immune cells, the training dataset ranked cell types in the same order as s-LDSC with the strongest association for monocytes, whereas the smaller test dataset showed a different pattern (Table 1).

**Table 1.**
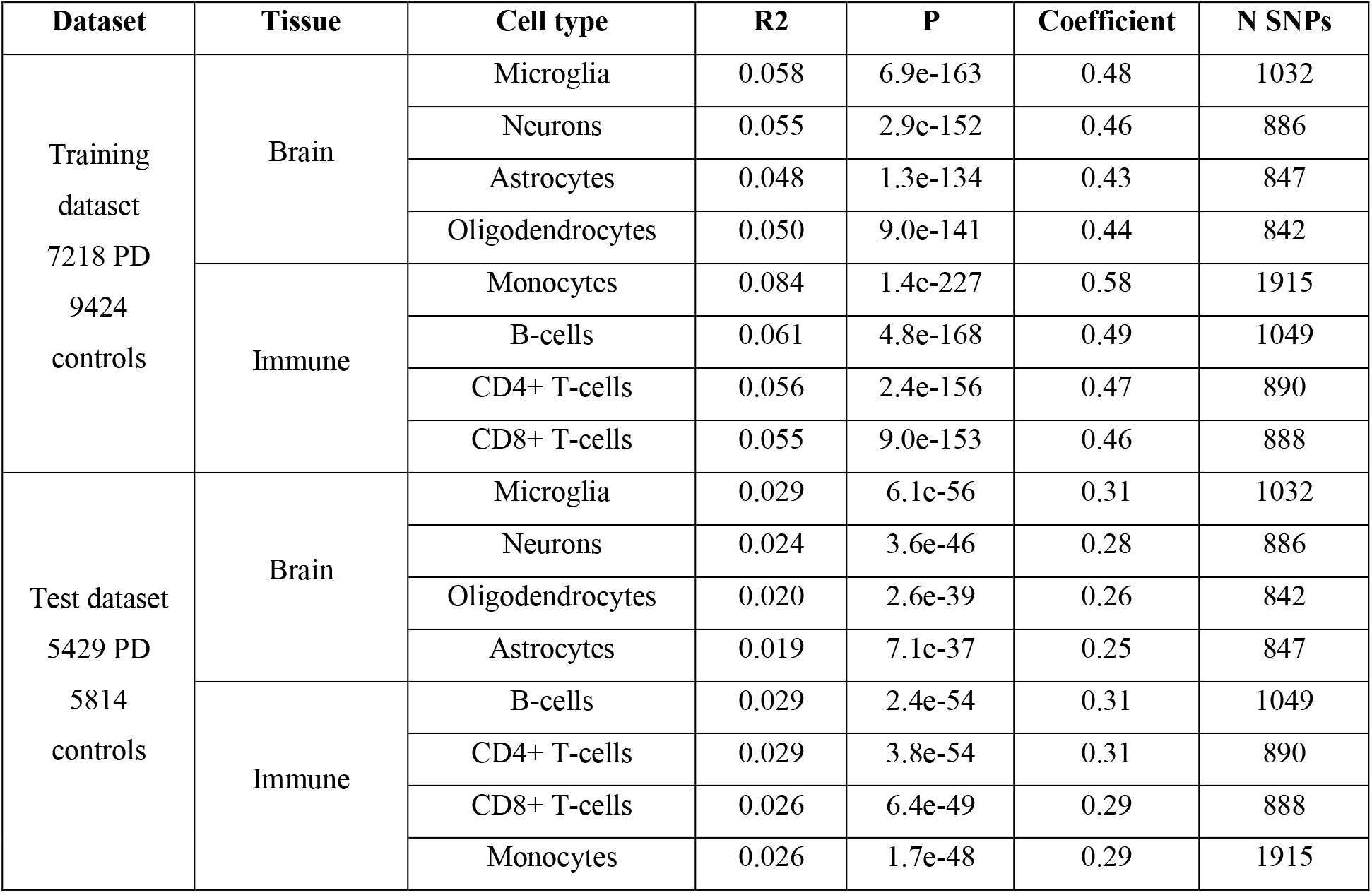
Polygenic risk score results stratified by brain cell type. Association results of polygenic risk scores modeled on summary statistics from an independent genome-wide association study and stratified to include variants within open chromatin regions for various cell types. Scores were generated using the PRSice software tool with a cutoff of p < 0.05 in the reference data. Within each dataset and tissue, results are ordered with the best performing model first.

### Nominating the microglial signature gene P2RY12 as driver of a specific GWAS signal in PD

Having shown microglial enrichment of common variant heritability based on genome-wide summary statistics, we explored whether available data could support examples of specific significant GWAS loci being driven by gene regulation in microglia. The estimated 95% credible sets included 1665 SNPs in total across 71 GWAS loci, out of which 33 SNPs across 22 loci overlapped with open chromatin in microglia, as compared to 21 SNPs across 16 loci in neurons. At 10 loci, such overlap was observed for microglia only, and no other brain cell types.

Gosselin *et al*. identified 881 transcripts comprising microglial signature in the human brain, using a cutoff of 10-fold increased expression relative to cortex tissue.^28^ They further intersected this microglial signature set with genes reported in previous studies as showing significant differential brain mRNA expression in neurodegenerative disease. Listed among 82 microglial signature genes identified as differentially expressed in PD brains by RNA sequencing^35^, is *P2RY12*, which is located close to a PD GWAS signal on chromosome 3. Credible set SNPs at this locus overlap with microglial, but not neuronal, open chromatin, located upstream of *P2RY12* (Figure 5).

**Figure 5.**
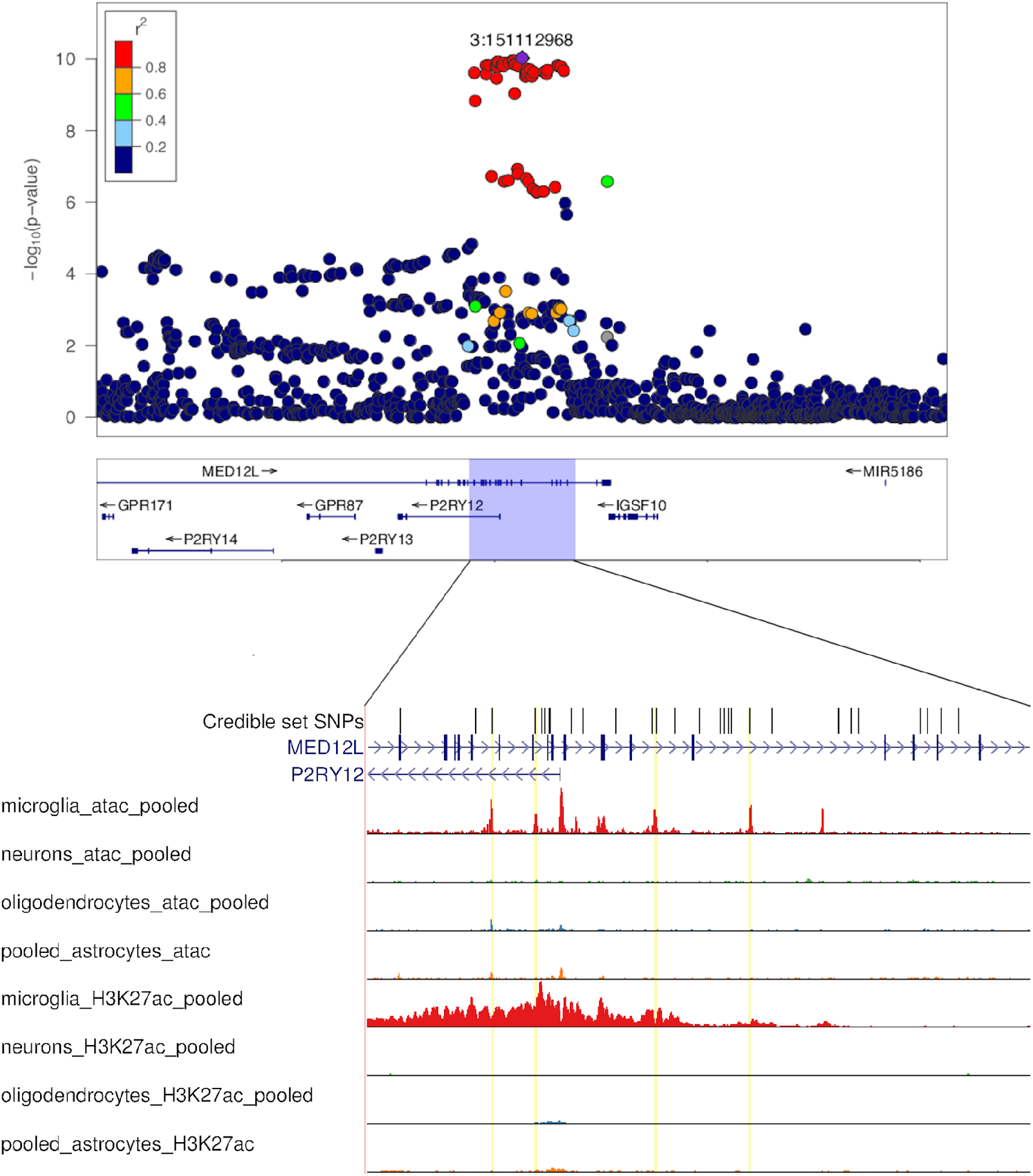
Overview of the *P2RY12* locus. The top panel shows a regional Manhattan plot of the *MED12L*/*P2RY12* locus in PD based on summary statistics from the most recent meta-analysis of PD GWAS.^9^ Below, the region containing significant SNPs is expanded to show the location of SNPs belonging to the 95% credible set in relation to brain cell specific results from ATAC-seq and Chromatin immunoprecipitation (ChIP)-seq of H3K27ac, a histone modificaion marking active enhancers. Credible set variants that co-localize with ATAC-seq peaks within a microglia-specific enhancer region are highlighted with yellow lines.

The expression profile across all tissues included in GTEx demonstrates that *P2RY12* is primarily expressed in brain tissue. We queried the Brain xQTLserve browser^27^ and found that the top-hit GWAS SNP rs11707416 was associated with *P2RY12* expression at p=1.2e-4, with no other gene reaching nominal significance. The strongest credible set SNP overlapping with microglia open chromatin, rs17283010 (r^2^ = 1 with rs11707416 in 1000 Genomes EUR), showed association with expression levels at p=1.5e-5. Similar results were found in the BRAINEAC database^30^ (rs11707416 eQTL for P2RY12 at p = 2.3e-4 averaged across brain regions) and PsychENCODE data^31^ (rs11707416 eQTL for P2RY12 at p = 1.2e-6) (see online resources). A PD GWAS locus browser was launched in 2020, summarizing the evidence supporting causality for specific genes at significant GWAS loci.^36^ The article presenting the browser also highlighted *P2RY12* as an example of a strong functional candidate, top ranked in the locus based on brain^29, 31^ and blood^37^ eQTL data, with a correlation between p-values for PD risk and *P2RY12* expression (Pearson correlation coefficient from 0.50 to 0.80 for the referenced eQTL datasets). Co-localization with regulatory regions of open chromatin was however not considered in this study.

## Discussion

In this study, we aimed to shed light on immune mechanisms in PD pathogenesis by investigating common variant heritability enrichment for specific cell types in the human brain and immune system. We observed significant enrichment in open chromatin regions of microglia, stronger than for any other class of brain cells. Although a role for microglia in pathogenesis is well established based on other lines of evidence, this finding is important in suggesting microglial dysregulation as a primary contributing cause of PD, not merely a secondary downstream phenomenon.

Microglial enrichment of heritability has been demonstrated in several studies of AD^12^ but has to our knowledge not been reported previously in PD. Reynolds *et al*. applied s-LDSC to PD summary statistics in microglia and other brain cells, but found no significant enrichment.^32^ A recent large-scale unbiased analysis taking a PRS approach to functional partitioning of genetic PD risk highlighted immune related processes as major contributors to PD etiology but did not report a specific enrichment for microglia.^14^ Common to both these studies, however, is that annotation was gene-centered. Our comparison of expression-based and open chromatin-based annotation approaches suggests that closing in on cell-specific regulatory DNA may provide higher sensitivity to detect enrichment in immune cells. Indeed, the p-value we observed for PRS-based enrichment analyses in the present work would have passed the significance threshold in the pathway-study published by Bandres-Ciga *et al*.^14^ The ATAC-seq data we used on brain cell types was generated as part of a previous study that also included s-LDSC analyses of PD.^18^ Similarly, the combination of single-cell ATAC-seq and s-LDSC was applied in a very recent study of PD and AD.^33^ Both these studies highlighted microglial enrichment in AD, yet reported no significant heritability enrichment for any brain cell type in PD. We note, however, that both analyses used summary statistics originating from a smaller sample set than we included here, and included more GWAS studies and cell-types resulting in a substantially higher multiple testing burden.

In addition to the significant finding observed for microglia, we detected an equally strong enrichment of heritability in monocyte open chromatin regions, the only immune cell type to pass an adjusted significance threshold. Several lines of evidence implicate both the innate and adaptive immune system in PD, recently reviewed by Tan *et al*.^2^ A study based on analyses of expression quantitative trait loci (eQTL) across leukocyte populations has proposed that while the adaptive immune system dominates in autoimmune disorders, innate immune mechanisms are of primary importance in neurodegenerative disorders, such as AD and PD.^5^ In line with this hypothesis, one study created composite annotations for different immune cell types, incorporating both gene expression and epigenetic data, and found the strongest immune cell enrichment signal for PD in monocytes.^11^ PD loci have also been associated with monocyte eQTLs in a transcriptome-wide association study.^13^ In contrast, others have applied s-LDSC combined with gene expression and epigenetic data and observed enrichment for most classes of immune cells in AD, yet only T-cells passing the significance threshold in PD.^3^

Microglia and monocytes both belong to the myeloid lineage, and we note a substantial overlap in their open chromatin regions as identified by ATAC-seq. This complicates the interpretation of our findings. Jointly analyzing heritability enrichment for microglia and monocytes in the same model, we found that only the microglial signal remained significant. Importantly, all types of genetic enrichment studies depend on experimentally generated data to annotate the genome, which for many years have been far more available for different classes of blood cells than for individual brain cell types. With functional genomic regions of monocytes largely overlapping those of microglia, there is a possibility that similarity to microglia could have been a driving factor behind some of the previously reported results linking monocytes to neurodegeneration.

An improved understanding of the genetic contribution to neuroinflammation in PD must not only highlight cell types, but also close in on specific risk loci to identify relevant genes and molecular mechanisms. Intersecting credible set SNPs from GWAS with cell-specific annotations and eQTL data we highlight *P2RY12* as a likely microglial gene driving a PD association signal, in line with a recent article presenting the PD GWAS browser.^36^ Very shortly before submission of our article, a preprint (not yet peer reviewed) was published mapping microglia-specific eQTLs to brain disorders, also nominating *P2RY12* in PD.^38^ *P2RY12* encodes the ADP receptor purinergic receptor P2Y, G protein– coupled 12, whose expression is often reported in the neuroscience literature as a marker of homeostatic, non-activated microglia.^39^ Disease-associated activation of microglia is associated with downregulation of *P2RY12*, coupled with upregulation of other genes, including the strongly AD-associated risk gene *TREM2*.^40-42^ Evidence indicates that *P2RY12* serves as a chemotactic receptor, directing movement of microglial cell processes toward sites of injury to enable repair, including maintenance of the blood-brain barrier.^43^ Interestingly, *P2RY12* is also the target molecule of the anti-thrombotic compound clopidogrel, which has been shown experimentally to inhibit microglial *P2RY12* in the context of blood-brain barrier breakdown.^44^ Further studies are clearly needed, but given that fact that this molecule is an established drug-target, we anticipate potentially important clinical implications if more evidence can be provided to support a causal role for *P2RY12* in PD.

Genetic evidence supporting a role for microglia have been abundant in AD.^12, 18, 33^ Although our study links common susceptibility variants to microglia also in PD, the genes and mechanisms involved are likely to be distinct, as previous studies have shown very limited genetic correlation between PD and AD.^45^ An important future challenge will be to disentangle how pathogenic microglial responses differ across different neurodegenerative disorders.

We note that the present study has several limitations. Our narrow focus on open chromatin in the cell type specific analyses prioritizes regulatory DNA while leaving out other classes of functionally relevant polymorphisms, such as coding variants and variants affecting splicing. Furthermore, the applied methodology captures only common variation. We therefore emphasize that our findings do not represent any attempt to provide a complete or final picture of the relative importance of different cell types for heritable PD risk. Furthermore, we found several sources of data to be compatible with the hypothesis that a PD GWAS locus on chromosome 3 is driven by an effect on *P2RY12* regulation, adding up to a compelling picture. Our study cannot, however, exclude other theoretically possible mechanisms, including variants in LD affecting a different gene through splicing or coding mechanisms, or effects involving other cell types than those studied here. Only functional experiments can ultimately resolve the mechanisms underlying typical GWAS signals in complex disorders.

In conclusion, we studied common variant PD heritability stratified for brain and immune cell types and found significant enrichment in microglia and monocytes. These results add to the growing body of evidence for immune mechanisms in PD pathogenesis and highlight microglia dysregulation as a primary contributing etiological factor. We also nominate the key microglial gene and clopidogrel target *P2RY12* as the likely driver of a PD GWAS signal. We anticipate further research into immune mechanisms and microglial function in neurodegeneration, ultimately aiming to develop targeted strategies to modulate these processes and develop novel effective therapies for PD.

## Data Availability

Summary statistics for the dataset excluding participants from 23andMe are available at https://bit.ly/2ofzGrk. The full GWAS summary statistics for the 23andMe discovery data set will be made available through 23andMe to qualified researchers under an agreement with 23andMe that protects the privacy of the 23andMe participants. Please visit https://research.23andme.com/dataset-access/ for more information and to apply to access the data.

## Acknowledgements

The GWAS summary statistics used in this study were generated in a meta-analysis including data from 23andMe, Inc. We would like to thank the research participants and employees of 23andMe for making this work possible. The full GWAS summary statistics for the 23andMe discovery data set will be made available through 23andMe to qualified researchers under an agreement with 23andMe that protects the privacy of the 23andMe participants. Please visit https://research.23andme.com/dataset-access/ for more information and to apply to access the data. MSA was funded by a grant from the South-Eastern Regional Health Authority, Norway. LP is supported by the Norwegian Health association. JH was supported by the UK Dementia Research Institute which receives its funding from DRI Ltd, funded by the UK Medical Research Council, Alzheimer’s Society and Alzheimer’s Research UK. JH is also supported by the · Medical Research Council (award number MR/N026004/1), Wellcome Trust Hardy (award number 202903/Z/16/Z), the Dolby Family Fund, National Institute for Health Research University College London Hospitals Biomedical Research Centre and BRCNIHR Biomedical Research Centre at University College London Hospitals NHS Foundation Trust and University College London.

## Author contributions

Conception and design of the study: LP Acquisition, analysis and/or interpretation of data: MSA, SBC, RHR, JH, MR, LK, ZGO, IH, IPDGC. Drafting of the manuscript and figures: MSA, LP. All authors contributed to critical revision of the manuscript.

## Online resources and publicly available data

23andMe: https://research.23andme.com/dataset-access/

Alkes group repository: https://alkesgroup.broadinstitute.org/LDSCORE/

BioMart: http://grch37.ensembl.org/biomart

Brain xQTLserve browser: http://mostafavilab.stat.ubc.ca/xqtl/

BRAINEAC: www.braineac.org

Genotype-Tissue Expression (GTEx): www.gtexportal.org

Linkage disequilibrium score regression: https://github.com/bulik/ldsc

LocusZoom: locuszoom.org/

MACS2: https://github.com/macs3-project/MACS/tree/master/MACS2

PAINTOR: https://github.com/gkichaev/PAINTOR_V3.0/wiki

PD GWAS locus browser: https://pdgenetics.shinyapps.io/GWASBrowser/

PRSice: https://www.prsice.info/

PsycENCODE: www.nimhgenetics.org/resources/psychencode

UCSC Genome Browser: https://genome.ucsc.edu

